# Performance of plasma amyloid, tau, and astrocyte biomarkers to identify cerebral AD pathophysiology

**DOI:** 10.1101/2022.02.21.22271198

**Authors:** Pâmela C. L Ferreira, Cécile Tissot, João Pedro Ferrari-Souza, Wagner S. Brum, Bruna Bellaver, Douglas T. Leffa, Joseph Therriault, Andréa L. Benedet, Firoza Z. Lussier, Mira Chamoun, Gleb Bezgin, Stijn Servaes, Jenna Stevenson, Nesrine Rahmouni, Vanessa Pallen, Min Su Kang, Nina Margherita Poltronetti, Dana L. Tudorascu, William E. Klunk, Victor L. Villemagne, Annie Cohen, Serge Gauthier, Eduardo R. Zimmer, Nicholas J. Ashton, Henrik Zetterberg, Kaj Blennow, Thomas K. Karikari, Pedro Rosa-Neto, Tharick A. Pascoal

## Abstract

**Introduction:** Plasma amyloid-β (Aβ), phosphorylated tau (p-tau), and glial fibrillar acid protein (GFAP) can identify Alzheimer’s disease (AD) pathophysiology with high accuracy. However, comparing their performance in the same individuals remains under-explored.

**Methods:** We compared the predictive performance of plasma Aβ42/40, p-tau(at threonine 181 and 231), neurofilament light (NfL), and GFAP to identify Aβ- and tau-PET positivity in 138 cognitive unimpaired (CU) and 87 cognitive impaired (CI) individuals.

**Results:** In CU, plasma p-tau231 had the best performance to identify both Aβ- and tau-PET positivity. In CI, plasma GFAP showed the best predictive accuracy to identify both Aβ and tau-PET positivity.

**Discussion:** Our results support plasma p-tau231 as a marker of early AD pathology and, that GFAP best identifies both PET Aβ and tau abnormalities in the brain of CI individuals. These findings highlight that the performance of blood-based protein biomarkers to identify the presence of AD pathophysiology is disease-stage dependent.

## 1. Introduction

The pathological hallmarks of Alzheimer’s disease (AD) are brain deposits of amyloid-β (Aβ) plaques and tau neurofibrillary tangles [1]. Measuring Aβ and tau levels in living subjects can increase clinicians’ diagnostic accuracy when assessing cognitively impaired (CI) patients [2] and can inform the risk of progression to dementia in cognitively unimpaired (CU) individuals [3]. Reduction of Aβ42/40 and increased tau [both total (t-tau) and phosphorylated tau (p-tau)] levels are the cerebrospinal fluid (CSF) biomarker signature of AD [4]. Similarly, positron emission tomography (PET) imaging can also be used for the visual identification and quantification of Aβ [5] and tau [6] aggregates. Although Aβ and tau can be accurately identified by CSF and PET [4, 7], these biomarker modalities’ costs, accessibility and relative invasiveness limit their use in clinical practice and trials. Thus, there is an urgent need for accessible and cost-effective blood methods to accurately identify these key AD processes. In fact, several studies demonstrated that Aβ, p-tau, neurofilament light (NfL), and glial fibrillary acidic protein (GFAP) levels are highly associated with the presence of AD hallmark proteinopathies in the human brain [8-19].

Plasma Aβ can be quantified by immunoprecipitation coupled to mass spectrometry (IP-MS) and by novel ultrasensitive immunoassay techniques [20-23], including fully automated clinical chemistry instruments [24]. Both methods can detect with relatively high precision individuals with abnormal brain Aβ burden or those at high risk of future conversion to Aβ-PET positivity [11, 20, 21, 23, 25]. More recently, plasma levels of tau protein phosphorylated at threonine 181 (p-tau181), 217 (p-tau217), and 231 (p-tau231) have shown high accuracy to detect both tau and Aβ pathologies [8, 11, 13]. Analysis of these three p-tau epitopes has shown comparable diagnostic accuracy for AD dementia, whereas p-tau-217 and p-tau231 has been suggested as a marker of early AD pathology [8, 11, 13, 26]. Although p-tau epitopes directly measure tau pathology, the fact that some epitopes become hyperphosphorylated in response to initial Aβ aggregation [27] suggesting that these markers would be more accurate than plasma Aβ to detect brain amyloidosis [8, 13]. NfL is a well-validated biomarker for tracking neurodegeneration and neuronal injury that has been shown to correlate with Aβ and tau but is not specific to AD, as abnormal levels are also described in several other neurodegenerative disorders [19, 28-31]. Studies have recently shown that plasma GFAP, a marker of astrocyte reactivity, can detect with high accuracy Aβ-PET positivity [10, 16, 32]. The high correlation has been attributed to the fact that astrocytes may show increased expression of GFAP in regions surrounding Aβ plaques [14, 33]. Although several studies have shown a strong correlation between the markers mentioned above and brain Aβ and tau levels, no previous study has directly compared the plasma performance of all these markers in the same individuals.

Here, we compared the performance of plasma Aβ, p-tau, GFAP, and NfL alone and in combination with demographics to identify both brain Aβ and tau pathology measured with PET in CU and CI individuals. We hypothesize that the performance of each biomarker will depend on the patient’s disease clinical stage.

## 2. Methods

### 2.1 Study participants

We assessed 225 individuals [138 CU elderly adults, 53 mild cognitive impairment (MCI) subjects and 34 AD dementia patients] from the Translational Biomarkers of Aging and Dementia (TRIAD) cohort of McGill University, Canada [34]. The participants underwent clinical and neuropsychological assessments, including Mini-Mental State Examination (MMSE) and the Clinical Dementia Rating (CDR). CU participants had a CDR score of 0 and no objective cognitive impairment. Participants with Mild cognitive impairment (MCI) had subjective and objective cognitive impairments CDR score of 0.5, and preserved activities of daily living. Alzheimer’s disease patients had a CDR score between 0.5 and 2 and met the National Institute on Aging and the Alzheimer’s Association criteria for probable Alzheimer’s disease determined by a physician [35]. For this study, individuals with a diagnosis of AD or MCI were classified as CI. Details on the information gathered from participants can be found here: https://triad.tnl-mcgill.com/.

### 2.2 Neuroimaging

Study participants had magnetic resonance imaging 3D T1-weighted MRI (3 T Siemens), tau [^18^F]MK6240 PET and Aβ [^18^F]AZD4694 PET scans with a brain-dedicated Siemens High Resolution Research Tomograph at the Montreal Neurological Institute. The acquisition and processing of the images followed standard protocols [34]. Braak stages were calculated according to previously described methods [36]. We considered individuals tau-PET positive (T+) if they were Braak stage II or above. Global [^18^F]AZD4694 standardized uptake value ratio (SUVR) value was estimated from the precuneus, prefrontal, orbitofrontal, parietal, temporal, anterior and posterior cingulate cortices [37]. We used the published [^18^F]AZD4694 cut-off value of 1.55 global SUVR [34] to classify the participants as Aβ positive (A+) or Aβ negative (A-).

### 2.3 Plasma measurements

Plasma p-tau181, p-tau231 and NfL were measured using in-house Single molecule array (Simoa) methods on an HD-X instrument (Quanterix, Billerica, MA, USA) at the Clinical Neurochemistry Laboratory, University of Gothenburg, Mölndal, Sweden [8, 38, 39]. Plasma GFAP was measured by the Simoa method using the commercial single-plex assay (No. 102336), while the plasma Aβ42, Aβ40 were quantified using a multiplexed assay. In addition, plasma Aβ42 and Aβ40 were measured using an IP-MS method with a previously described method [40].

### 2.4 Statistical analysis

Statistical analyses were performed using R statistical software version 4.0.5 (http://www.r-project.org/). The demographic characteristics considered in the statical analysis were sex, age, and apolipoprotein E4 (APOE ε4) carriage status. Voxel-wise statistics were conducted using MATLAB software version 9.2 (http://www.mathworks.com) with the VoxelStats package [41]. Descriptive statistics, including means and standard deviations, were calculated for CU and CI groups, and comparisons were performed using Student t-tests for continuous variables and chi-square tests for the categorical ones. The association between the plasma biomarkers was assessed using Pearson correlations and linear regressions. In the ROI-based multiple regression models, partial residuals generated with the R function termplot were used to graphically represent the investigated associations [42]. Model goodness-of-fit was evaluated using R-square analyses. Building from basic demographics including age, sex, and APOE ε4 genotype as predictors, we sequentially added plasma biomarkers in all possible combinations and evaluated models’ performance alone and in combination with demographics. We assessed and compared the discriminative performances of each model using receiver operating characteristic (ROC) and area under the curve (AUC) with the DeLong test. Statistical significance was set as p<0.05 after considering Bonferroni correction for multiple comparison tests.

## 3. Results

### 3.1 Participant Characteristics and Biomarker profile

In the CU group, 23.1% of individuals were A+ and 17.0% were T+. In the CI group, 81.0% were Aβ+ and 72.0 % were Tau+. The concentrations of p-tau231 and GFAP were 1.5-fold higher in the CI compared to the CU group (P < 0.0001). Similarly, p-tau181 concentrations were 1.7-fold higher in the CI group (P < 0.0001) and NfL concentrations were 1.2-fold higher in the CI group (P = 0.01). Simoa and IP-MS Aβ42/40 were not significantly different between diagnostic groups. We also compared the groups regarding their A and T status, and the results remain similar. Demographic and biomarkers characteristics of the population are summarized in Table 1.

**Table 1.** Demographics and characteristics of the population

|  | CU | CI |  | CU A- | CU A+ |  | CI A- | CI A+ |  | CU T- | CU T+ |  | CI T- | CI T+ |  |
| --- | --- | --- | --- | --- | --- | --- | --- | --- | --- | --- | --- | --- | --- | --- | --- |
|  | (N=138) | (N=87) | P value | (N=99) | (N=41) | P value | (N=19) | (N=72) | P value | (N=99) | (N=41) | P value | (N=19) | (N=72) | P value |
| **Demographic characteristics** |  |  |  |  |  |  |  |  |  |  |  |  |  |  |  |
| Age, y<sup>a</sup> | 71.7 (5.93) | 69.8 (7.30) | 0.06 | 70.7 (5.77) | 75.4 (5.24) | < 0.0001 | 70.3 (6.97) | 69.5 (7.16) | 0.651 | 71.1 (6.11) | 73.4 (5.09) | 0.02 | 70.5 (7.65) | 69.2 (7.49) | 0.493 |
| Education, y<sup>a</sup> | 15.4 (3.69) | 14.8 (3.53) | 0.346 | 15.6 (3.73) | 14.9 (3.28) | 0.210 | 14.0 (3.71) | 14.9 (3.29) | 0.323 | 15.2 (3.56) | 15.7 (3.87) | 0.487 | 14.0 (3.45) | 15.0 (3.47) | 0.271 |
| Female, No. (%) | 95 (66.9) | 48.0 (54.5) | 0.756 | 99.0 (63.1%) | 34.0 (75.6%) | 0.167 | 11.0 (47.8%) | 57.0 (54.3%) | 0.028 | 63.0 (63.6%) | 29.0 (70.7%) | 0.542 | 11.0 (57.9%) | 38.0 (52.8%) | 0.889 |
| APOE ε4, number (%) | 39.0 (27.5) | 42.0 (47.7) | 0.002 | 39 (24.8%) | 15.0 (33.3%) | 0.345 | 5.0 (21.7%) | 54.0 (51.4%) | 0.74 | 23.0 (23.2%) | 15.0 (36.6%) | 0.159 | 2.0 (10.5%) | 42.0 (58.3%) | < 0.0001 |
| **Cognition** |  |  |  |  |  |  |  |  |  |  |  |  |  |  |  |
| MMSE<sup>a</sup> | 29.1 (1.18) | 24.9 (5.41) | < 0.0001 | 29.3 (1.12) | 29.1 (1.05) | 0.445 | 28.3 (1.49) | 23.6 (6.43) | < 0.0001 | 29.1 (1.09) | 29.1 (1.41) | 0.723 | 27.7 (1.59) | 23.8 (5.99) | < 0.0001 |
| **Plasma measures, pg/mL** |  |  |  |  |  |  |  |  |  |  |  |  |  |  |  |
| p-tau231<sup>a</sup> | 14.7 (7.35) | 22.1 (10.8) | < 0.0001 | 12.8 (5.74) | 23.4 (13.1) | < 0.0001 | 12.3 (7.14) | 24.1 (10.3) | < 0.0001 | 13.9 (7.28) | 18.5 (11.4) | 0.021 | 13.6 (7.33) | 24.5 (10.7) | < 0.0001 |
| p-tau181<sup>a</sup> | 10.8 (4.93) | 18.7 (8.89) | < 0.0001 | 9.9 (4.09) | 16.0 (12.1) | 0.002 | 12.0 (6.53) | 20.4 (9.45) | < 0.0001 | 10.4 (4.85) | 13.9 (10.6) | 0.05 | 13.4 (6.93) | 20.8 (9.78) | < 0.0001 |
| GFAP<sup>a</sup> | 215 (112) | 319.0 (170) | < 0.0001 | 187.0 (91.0) | 296.0 (129) | < 0.0001 | 149.0 (63.2) | 350.0 (160) | < 0.0001 | 210.0 (113) | 236.0 (112) | 0.226 | 190.0 (87.5) | 353.0 (168) | < 0.0001 |
| Aβ40 (Simoa)<sup>a</sup> | 254 (62.3) | 247.0 (56.5) | 0.321 | 225.8 (54.8) | 251.8 (52.3) | 0.07 | 250.6 (67.8) | 269.1 (60.6) | 0.908 | 221.2 (55.9) | 267.6 (45.91) | 0.02 | 243.6 (59.7) | 272.7 (62.6) | 0.586 |
| Aβ42 (Simoa)<sup>a</sup> | 8.99 (2.98) | 9.4 (3.07) | 0.213 | 10.3 (3.21) | 8.6 (3.21) | 0.113 | 9.5 (3.2) | 8.7 (2.93) | 0.404 | 9.5 (3.2) | 10.1 (2.93) | 0.647 | 8.8 (3.1) | 9 (3.0) | 0.903 |
| Aβ42/Aβ40 (Simoa)<sup>a</sup> | 0.05 (0.20) | 0.05 (0.19) | 0.26 | 0.08<br>(0.301) | 0.03 (0.01) | 0.09 | 0.04 (0.010) | 0.03<br>(0.010) | 0.219 | 0.07 (0.28) | 0.04 (0.01) | 0.306 | 0.03 (0.01) | 0.03<br>(0.009) | 0.644 |
| Aβ40 (IP-MS)<sup>a</sup> | 279.0 (44.4) | 279.0<br>(53.6) | 0.861 | 252.7<br>(48.4) | 279.7<br>(46.7) | 0.580 | 300.8 (55.1) | 277.1<br>(48.1) | 0.06 | 260.7<br>(47.7) | 278.9<br>(48.9) | 0.880 | 309.3<br>(64.3) | 278.1<br>(45.4) | 0.03 |
| Aβ42 (IP-MS)<sup>a</sup> | 29.9 (14.0) | 27.8 (13.7) | 0.382 | 29.7 (12.6) | 37.6 (36.2) | 0.07 | 29.4 (7.6) | 31.2 (14.8) | 0.679 | 30.2 (13.8) | 35.5 (30.6) | 0.188 | 27.1 (14.5) | 32.2 (14.6) | 0.814 |
| Aβ42/Aβ40 (IP-MS)<sup>a</sup> | 0.11 (0.05) | 0.11 (0.05) | 0.756 | 0.11 (0.05) | 0.104<br>(0.07) | 0.709 | 0.09 (0.02) | 0.102<br>(0.051) | 0.345 | 0.12 (0.05) | 0.11 (0.06) | 0.726 | 0.09 (0.02) | 0.11 (0.06) | 0.173 |
| NfL <sup>a</sup> | 25.0 (16.9) | 31.0 (13.2) | 0.01 | 22.2 (9.36) | 29.4 (22.1) | 0.042 | 23.9 (8.45) | 29.8 (12.1) | 0.008 | 22.2 (9.36) | 29.4 (22.1) | 0.09 | 23.9 (8.45) | 29.8 (12.1) | 0.07 |
| **Neuroimaging** |  |  |  |  |  |  |  |  |  |  |  |  |  |  |  |
| [<sup>18</sup>F]AZD4694 global<br>SUVR<sup>a</sup> | 1.6 (0.35) | 2.20 (0.59) | < 0.0001 | 1.3 (0.09) | 2.0 (0.341) | < 0.0001 | 1.3 (0.110) | 2.4 (0.451) | < 0.0001 | 1.4 (0.28) | 1.6 (0.47) | 0.009 | 1.5 (0.44) | 2.4 (0.48) | < 0.0001 |
| Aβ+ (%) | 23.0 | 81.0 | - | 0 | 100 | - | 0 | 100 | - | 0 | 100 | - | 0 | 100 | - |
| [<sup>18</sup>F]MK-6240 global<br>SUVR<sup>a</sup> | 1.1 (0.29) | 2.2 (0.99) | < 0.0001 | 0.99<br>(0.224) | 1.3 (0.478) | < 0.0001 | 0.95 (0.229) | 2.4 (0.837) | < 0.0001 | 0.94 (0.17) | 1.3 (0.42) | <<br>0.0001 | 0.93 (0.19) | 2.5 (0.83) | < 0.0001 |
| Tau + (%) | 17.0 | 72.0 | - | 0 | 100 | - | 0 | 100 | - | 0 | 100 | - | 0 | 100 | - |

P values indicate the results of the analysis of variance to assesses difference between groups except for gender and APOE ε4 status where a contingency chi-square was performed. <sup>a</sup>Mean (SD). CU = cognitively unimpaired, CI = cognitively impaired. Aβ = amyloid β. p-tau181 = tau phosphorylated at threonine 181, p-tau231 = tau phosphorylated at threonine 231, GFAP = glial fibrillary acidic protein, NfL = neurofilament light, MMSE Mini Mental State Examination. IP-MS = immunoprecipitation mass spectrometry.

### 3.2 Correlations Between Plasma Biomarkers

Pearson correlations coefficients between plasma biomarkers are shown in Figure 1. In the CU group, associations between plasma p-tau181 and p-tau231 measures (P < 0.0001), as well as GFAP with NfL (P < 0.0001) were statistically significant after multiple comparison correction. Aβ42/40 (Simoa and IP-MS) were not associated with each other or any other plasma marker. In the CI group, the correlation between p-tau, GFAP and NfL measures were significant (P < 0.0001). Aβ42/40 (Simoa/IP-MS) were not associated with each other or the other plasma markers.

**Figure 1.**
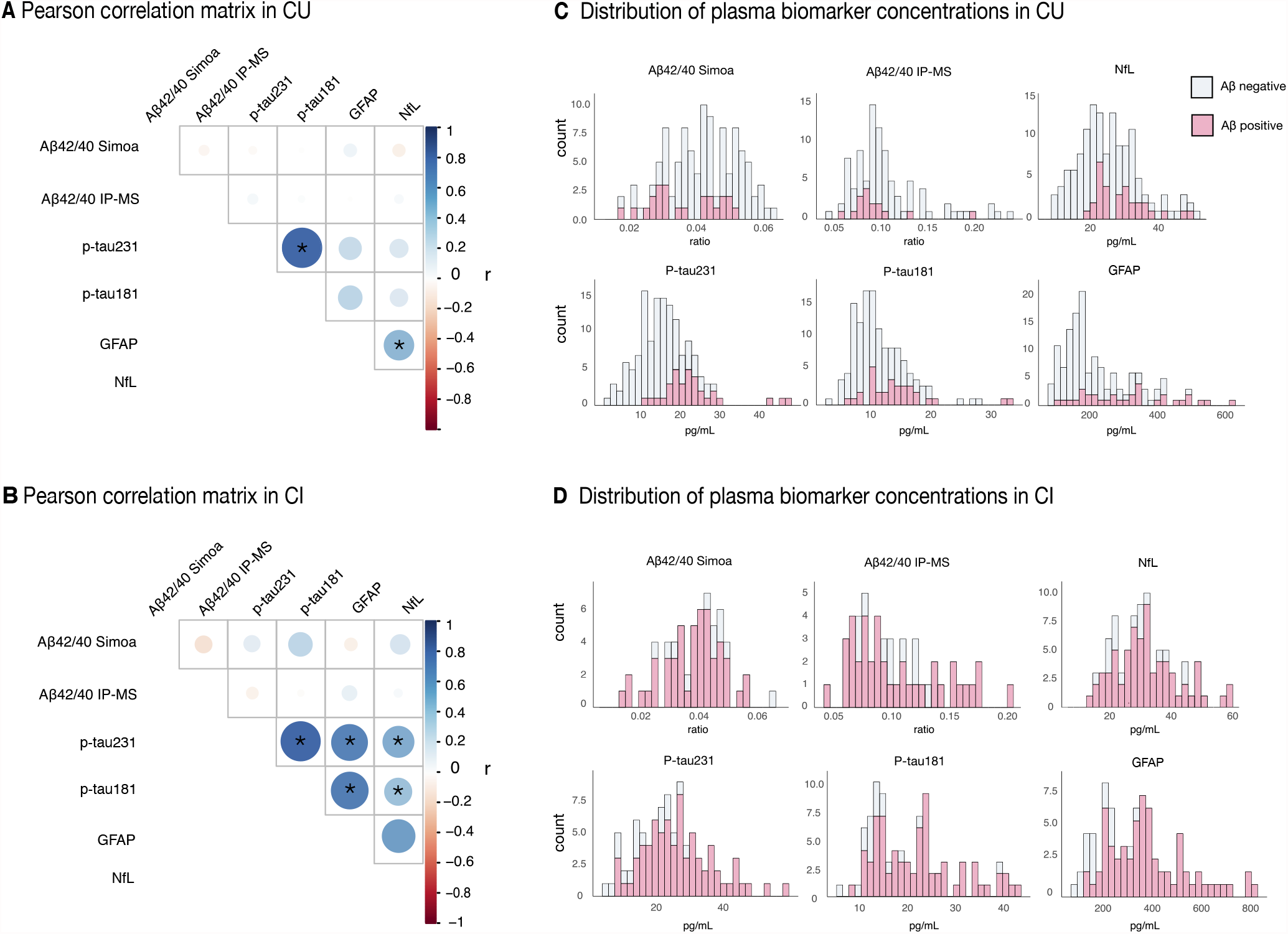
Correlation coefficients and distribution of plasma biomarkers in CU and CI individuals. The panel on the top show the Pearson coefficients in CU (A) and CI (B) groups. The panel on the middle show the histograms with the distribution of each biomarker regarding their Al3 status in CU (C) and CI (D) groups. * Indicates the Pearson correlation coefficients that survived to Bonferroni correction for multiple comparison in each clinical group (15 tests, significant P value < 0.003). CU = Cognitively unimpaired, CI = Cognitively impaired. Histograms with the distribution of each biomarker regarding their Tau status are presented in Supplemental Figure 1.

### 3.3 Voxel-wise association of Aβ-PET SUVR with plasma biomarkers

We performed voxel-wise linear regression between plasma markers and Aβ-PET SUVR in the CU and CI groups accounting for demographics. In the CU group, voxel-wise regression showed a significant positive association between Aβ-PET SUVR and plasma p-tau (p-tau231>p-tau181) and GFAP concentrations in the temporal and frontal cortices (Figure 2A). No significant voxel-wise associations were found between Aβ-PET and plasma Aβ42/40 (Simoa) and NfL concentrations. Aβ42/40 (IP-MS) concentrations showed significant positive, rather than negative, association with Aβ-PET SUVR in the temporal cortex (Supplemental Figure 2). In the CI group, voxel-wise regressions presented a significant positive association between Aβ-PET SUVR with plasma p-tau231 (but not p-tau181) and GFAP concentrations across the whole brain cortex after correction for multiple comparisons (Figure 2A). P-tau181 concentrations showed significant positive association with Aβ-PET in the precuneus and temporal cortex. No significant associations were observed between Aβ-PET with plasma Aβ42/40 measured by Simoa and IP-MS or NfL concentrations in CI individuals after multiple comparison corrections.

**Figure 2.**
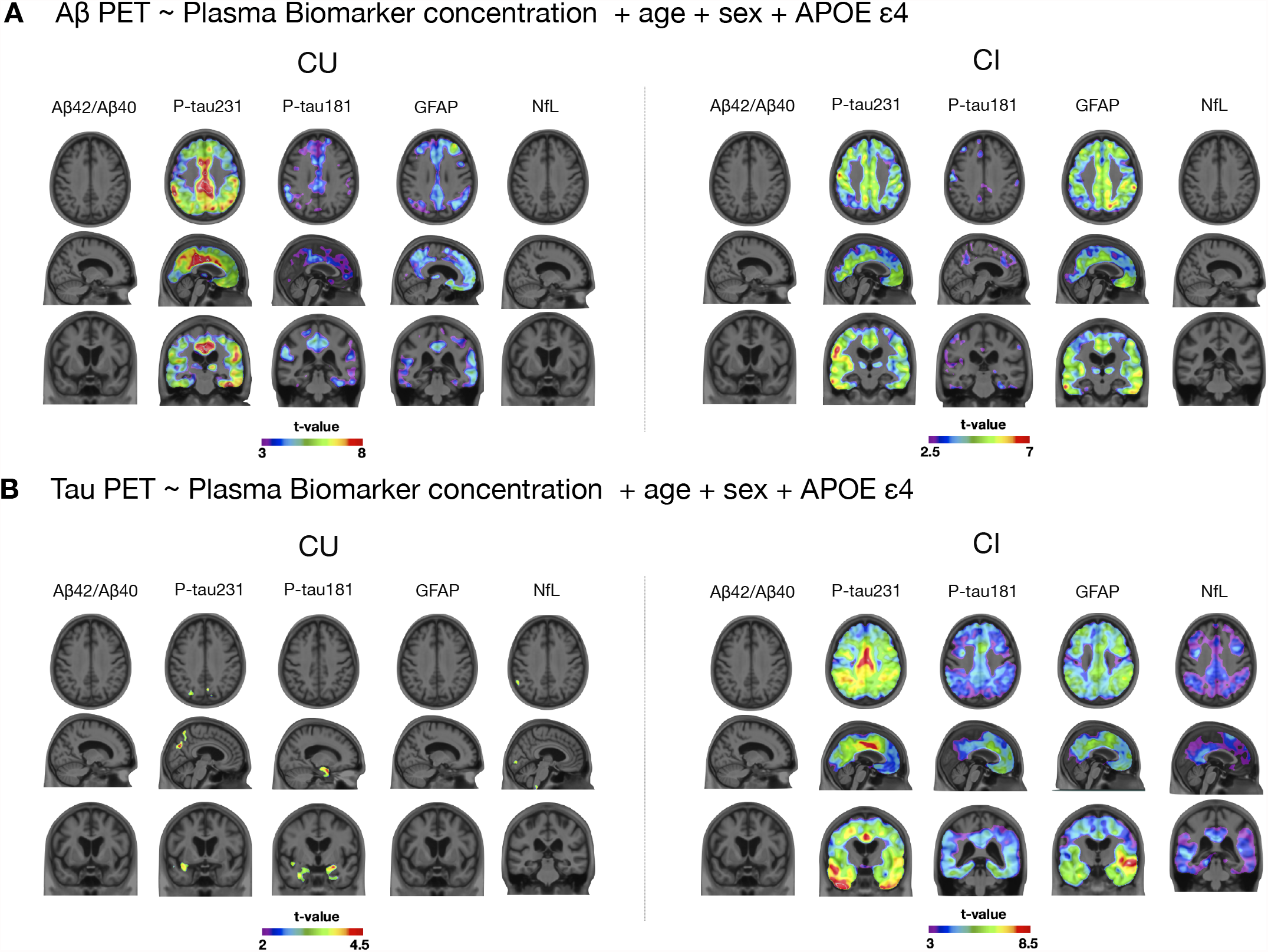
Voxel-wise associations of A [^18^F A 46 4 SUVR and tau [^18^F]MK-6240 SUVR with plasma biomarker concentrations in CU and CI individuals. The figure shows voxel-wise t-statistical maps of regressions between PET SUVR and plasma markers after RFT-correction for multiple comparisons at P < 0.05. Panel A shows the regions with a significant positive association between A [^18^F]AZD4694 and plasma markers in CU and CI individuals. No significant negative associations were found after multiple comparison correction. The panel B shows the regions indicating a significant positive association between tau [^18^F]MK-6240 SUVR and plasma markers in CU and CI individuals. No negative association was found between biomarkers after multiple comparison cor-rection. The covariates included in the voxel-wise linear regression models were age, sex and APOE ε4. A42/40 measure by Simoa. Cognitively unimpaired, Cl Cognitively impaired. Voxel-wise association of [^18^F A 46 4 SUVR and tau [^18^F MK-6240 SUVR with plasma A42/40 measured by lP-MS can be found in Supplementary Figure 2.

### 3.4 Voxel-wise association of tau-PET SUVR with plasma biomarkers

In the CU group, voxel-wise regression indicated a significant positive association between tau-PET SUVR and p-tau231 in small clusters in the precuneus and temporal cortices and p-tau181 in the entorhinal cortex, after correction for multiple comparisons (Figure 2B). No significant association was observed between Aβ-PET and plasma Aβ42/40 (Simoa/IP-MS), GFAP, and NfL concentrations in CU individuals. In the CI group, voxel-wise regressions indicated a significant positive association between tau-PET SUVR with plasma p-tau231 and GFAP concentrations across the brain cortex (Figure 2B). The p-tau181 and NfL concentrations showed widespread significant positive association with tau PET SUVR in the precuneus, frontal, and temporal cortices (P < 0.05). No significant association was found between tau-PET SUVR and plasma Aβ42/40 (measured by Simoa and IP-MS) in CI individuals.

### 3.5 ROI-wise association of Aβ-PET SUVR with plasma biomarkers

In the CU group, we found a significant positive association between p-tau231, p-tau181 and GFAP concentrations and global Aβ-PET SUVR values (Figure 3A). No significant association was found between global Aβ-PET SUVR values and Aβ42/40 (Simoa and IP-MS) and NfL values. The model using p-tau231 concentrations plus demographics as predictors explained the highest variability in the global Aβ-PET SUVR values (R-squared: 0.33). In the CI group, p-tau231, ptau181, and GFAP concentrations were significantly positive associated with global Aβ-PET SUVR. No significant association was found between global Aβ-PET SUVR and Aβ42/40 (Simoa and IP-MS) and NfL. P-tau231 concentration plus demographics explained the highest variability in the global Aβ-PET SUVR values (R-squared: 0.30), closely followed by GFAP plus demographics (R-squared: 0.28) (Supplemental Table 1). Additional analysis adding to the model tau-PET SUVR as a covariate was performed, and the association between Aβ-PET SUVR and p-tau231 and GFAP in the CU group remained significant. However, it was insignificant in the CI group (Supplemental Table 2).

**Figure 3.**
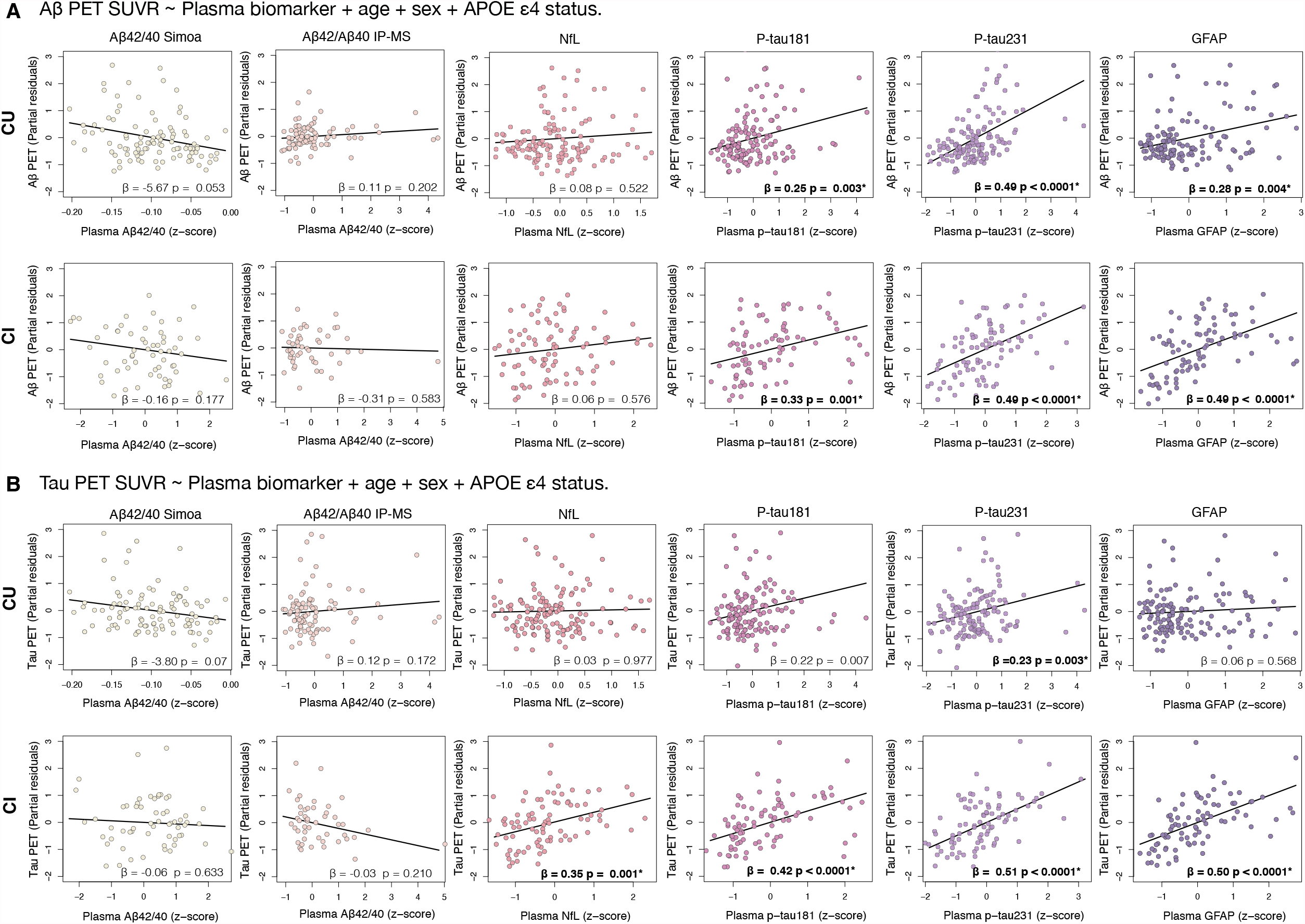
ROI-wise association of A0 [^18^F]AZD4694 SUVR and tau [^18^F]MK-6240 SUVR with plasma biomarker values. The plots show the partial residual of ROI-based linear regressions testing the association between PET SUVR values (global A β value and entorhinal tau value) corrected by age, sex and APOE s4 and plasma biomarker concentrations in CU (A) and Cl (B). ’Indicates the correlations that survived to Bonferroni correction for multiple comparison inside each clinical group (12 tests, corrected P value < 0.0042). CU = Cognitively unimpaired, Cl = Cognitively impaired.

### 3.6 ROI-wise association of tau-PET SUVR with plasma biomarkers

In the CU group, p-tau231 was significantly associated with entorhinal tau-PET SUVR. No significant association was found with Aβ42/40 (Simoa and IP-MS), p-tau181, GFAP, and NfL (Figure 3B). P-tau231 concentration plus demographics was the combination that best explains variability in the entorhinal tau-PET SUVR (R-squared: 0.15). In the CI group, entorhinal cortex SUVR was significantly associated with p-tau231, p-tau181, NfL, and GFAP. No significant association was found with Aβ42/40 (Simoa and IP-MS) values. P-tau231 plus demographics explained the highest variability in the entorhinal tau-PET SUVR (R-squared: 0.45), closely followed by GFAP plus demographics (R-squared: 0.44). Additional analysis adding to the model Aβ-PET SUVR as a covariate was performed, and the association between tau-PET SUVR and GFAP in the CI group remains significant while in the CU were not significant (Supplemental Table 2).

### 3.7 Prediction of Aβ-PET positivity with plasma biomarker concentrations

Plasma p-tau231 (alone or accounting for demographic characteristics) demonstrated higher performance in predicting A+ than demographics alone in the CU group (P < 0.0001; Figure 4A). On the other hand, Aβ42/40 (Simoa and IP-MS), p-tau181, GFAP, and NfL (alone or accounting for demographic characteristics) did not show a significantly better predictive performance than demographics alone (95% confidence interval range: 0.615 – 0.811). DeLong test comparisons showed that the model including p-tau231 and demographics (AUC = 87.7%) was significantly better predictor than models including the other plasma makers (Aβ42/40 (Simoa and IP-MS), p-tau181, GFAP, and NfL) alone (Table 2) or in combination with demographics (Supplemental Table 3). In the CI group, GFAP (alone or accounting for demographic characteristics) and p-tau231 (alone or accounting for demographic characteristics) showed higher performance than when only including demographics alone (P < 0.0001; Figure 4B) to detect Aβ positivity. On the other hand, Aβ42/40 (Simoa and IP-MS), p-tau181, and NfL, (alone or accounting for demographic characteristics) did not show a better predictive performance than demographics alone (95% confidence interval range: 0.551 – 0.793). DeLong test comparison showed that the model including GFAP plus demographics (AUC = 93.6%) was significantly in predicting A+ better than the Aβ42/40 (Simoa and IP-MS) p-tau231, p-tau181, and NfL alone (Table 2) or accounting for demographics (Supplemental Table 3).

**Table 2.**
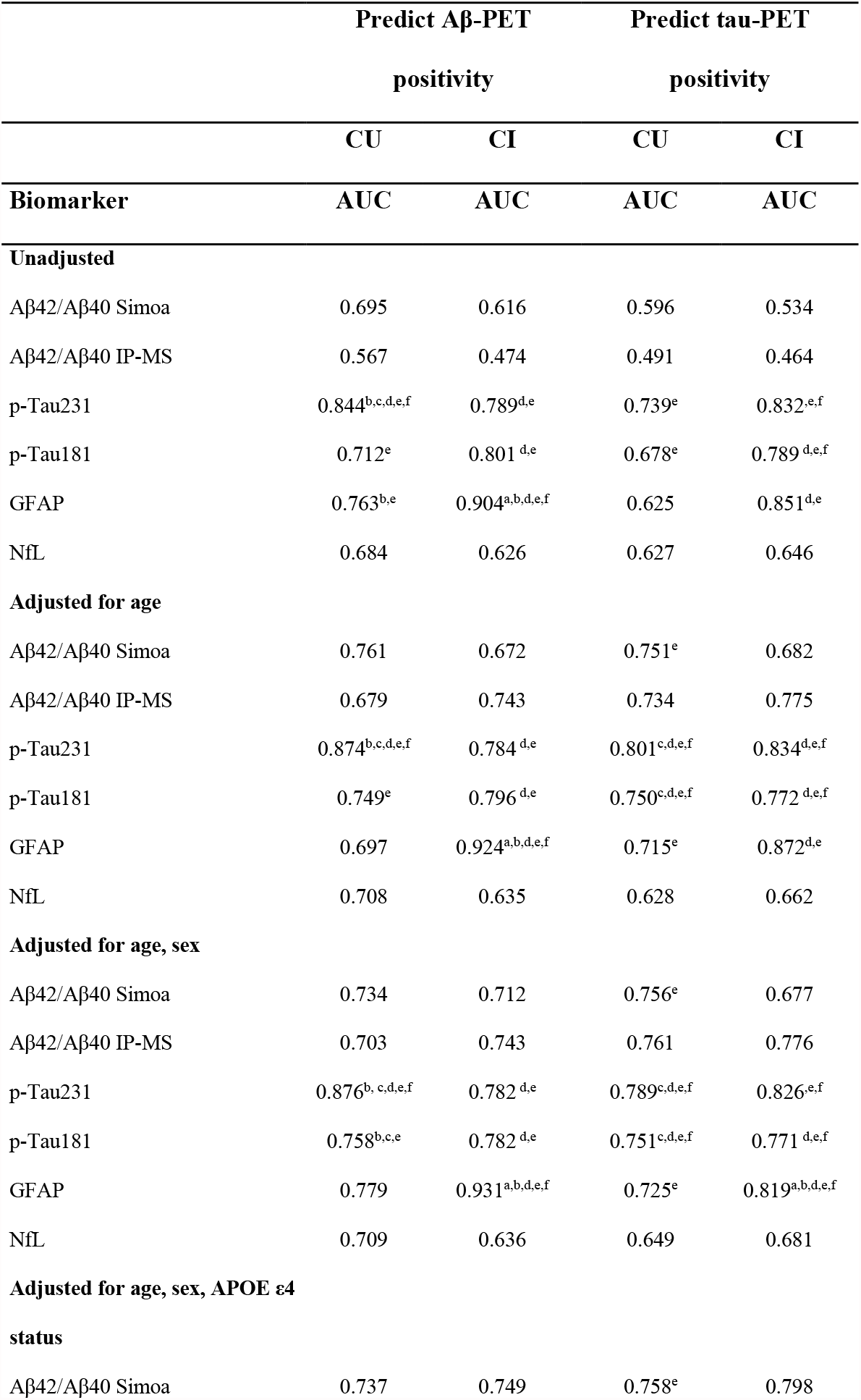
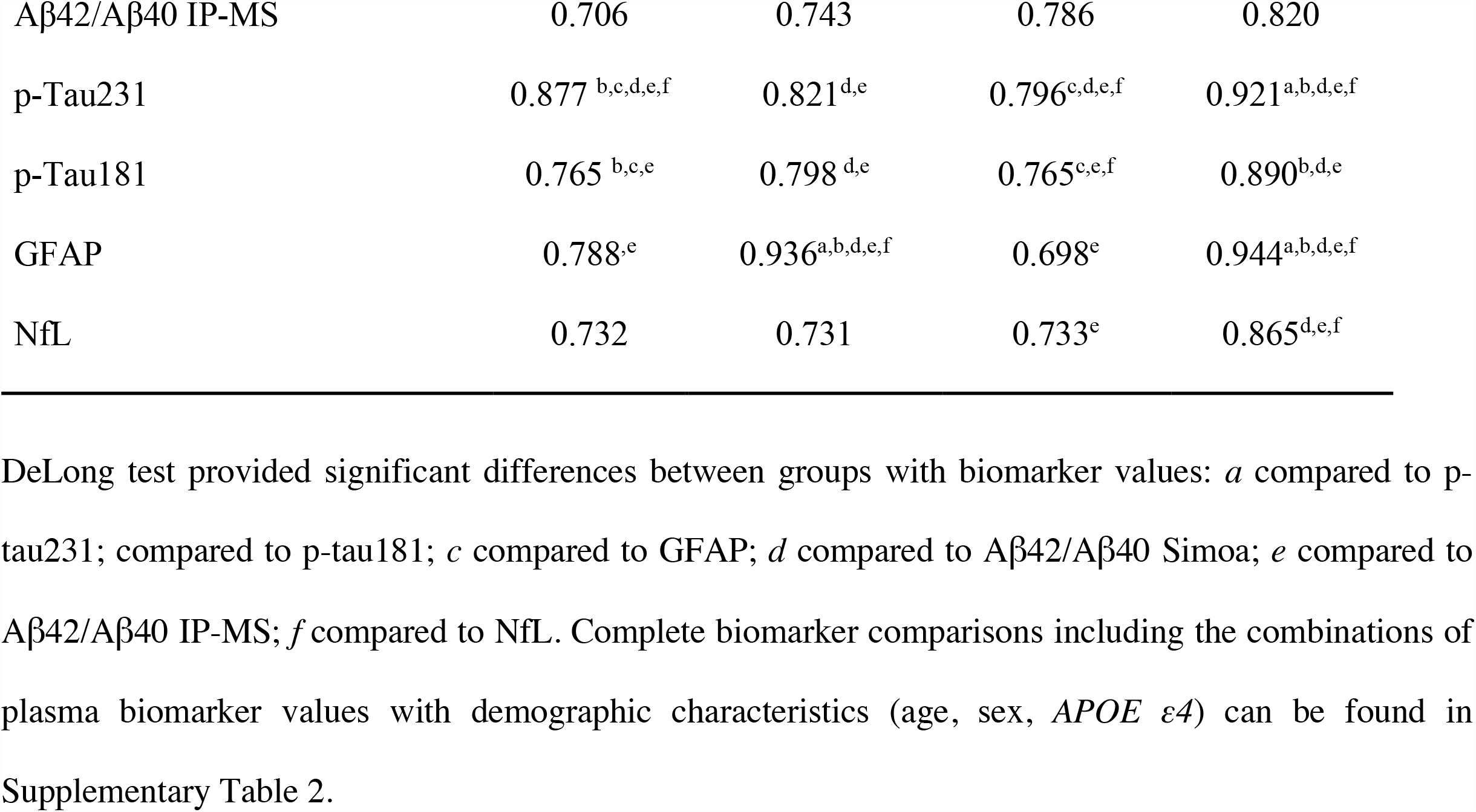
Performance of plasma biomarker to predict Aβ-PET and tau-PET positivity.

| Biomarker | Predict Aβ-PET | | Predict tau-PET | |
| --- | --- | --- | --- | --- |
|  | positivity |  | positivity |  |
|  | CU | CI | CU | CI |
|  | AUC | AUC | AUC | AUC |
| **Unadjusted** |  |  |  |  |
| Aβ42/Aβ40 Simoa | 0.695 | 0.616 | 0.596 | 0.534 |
| Aβ42/Aβ40 IP-MS | 0.567 | 0.474 | 0.491 | 0.464 |
| p-Tau231 | 0.844<sup>b,c,d,e,f</sup> | 0.789<sup>d,e</sup> | 0.739<sup>e</sup> | 0.832<sup>e,f</sup> |
| p-Tau181 | 0.712<sup>e</sup> | 0.801<sup>d,e</sup> | 0.678<sup>e</sup> | 0.789<sup>d,e,f</sup> |
| GFAP | 0.763<sup>b,e</sup> | 0.904<sup>a,b,d,e,f</sup> | 0.625 | 0.851<sup>d,e</sup> |
| NfL | 0.684 | 0.626 | 0.627 | 0.646 |
| **Adjusted for age** |  |  |  |  |
| Aβ42/Aβ40 Simoa | 0.761 | 0.672 | 0.751<sup>e</sup> | 0.682 |
| Aβ42/Aβ40 IP-MS | 0.679 | 0.743 | 0.734 | 0.775 |
| p-Tau231 | 0.874<sup>b,c,d,e,f</sup> | 0.784<sup>d,e</sup> | 0.801<sup>c,d,e,f</sup> | 0.834<sup>d,e,f</sup> |
| p-Tau181 | 0.749<sup>e</sup> | 0.796<sup>d,e</sup> | 0.750<sup>c,d,e,f</sup> | 0.772<sup>d,e,f</sup> |
| GFAP | 0.697 | 0.924<sup>a,b,d,e,f</sup> | 0.715<sup>e</sup> | 0.872<sup>d,e</sup> |
| NfL | 0.708 | 0.635 | 0.628 | 0.662 |
| **Adjusted for age, sex** |  |  |  |  |
| Aβ42/Aβ40 Simoa | 0.734 | 0.712 | 0.756<sup>e</sup> | 0.677 |
| Aβ42/Aβ40 IP-MS | 0.703 | 0.743 | 0.761 | 0.776 |
| p-Tau231 | 0.876<sup>b, c,d,e,f</sup> | 0.782<sup>d,e</sup> | 0.789<sup>c,d,e,f</sup> | 0.826<sup>e,f</sup> |
| p-Tau181 | 0.758<sup>b,c,e</sup> | 0.782<sup>d,e</sup> | 0.751<sup>c,d,e,f</sup> | 0.771<sup>d,e,f</sup> |
| GFAP | 0.779 | 0.931<sup>a,b,d,e,f</sup> | 0.725<sup>e</sup> | 0.819<sup>a,b,d,e,f</sup> |
| NfL | 0.709 | 0.636 | 0.649 | 0.681 |
| **Adjusted for age, sex, APOE ε4 status** |  |  |  |  |
| Aβ42/Aβ40 Simoa | 0.737 | 0.749 | 0.758<sup>e</sup> | 0.798 |
| Aβ42/Aβ40 IP-MS | 0.706 | 0.743 | 0.786 | 0.820 |
| p-Tau231 | 0.877<sup>b,c,d,e,f</sup> | 0.821<sup>d,e</sup> | 0.796<sup>c,d,e,f</sup> | 0.921<sup>a,b,d,e,f</sup> |
| p-Tau181 | 0.765<sup>b,c,e</sup> | 0.798<sup>d,e</sup> | 0.765<sup>c,e,f</sup> | 0.890<sup>b,d,e</sup> |
| GFAP | 0.788<sup>e</sup> | 0.936<sup>a,b,d,e,f</sup> | 0.698<sup>e</sup> | 0.944<sup>a,b,d,e,f</sup> |
| NfL | 0.732 | 0.731 | 0.733<sup>e</sup> | 0.865<sup>d,e,f</sup> |

**Figure 4.**
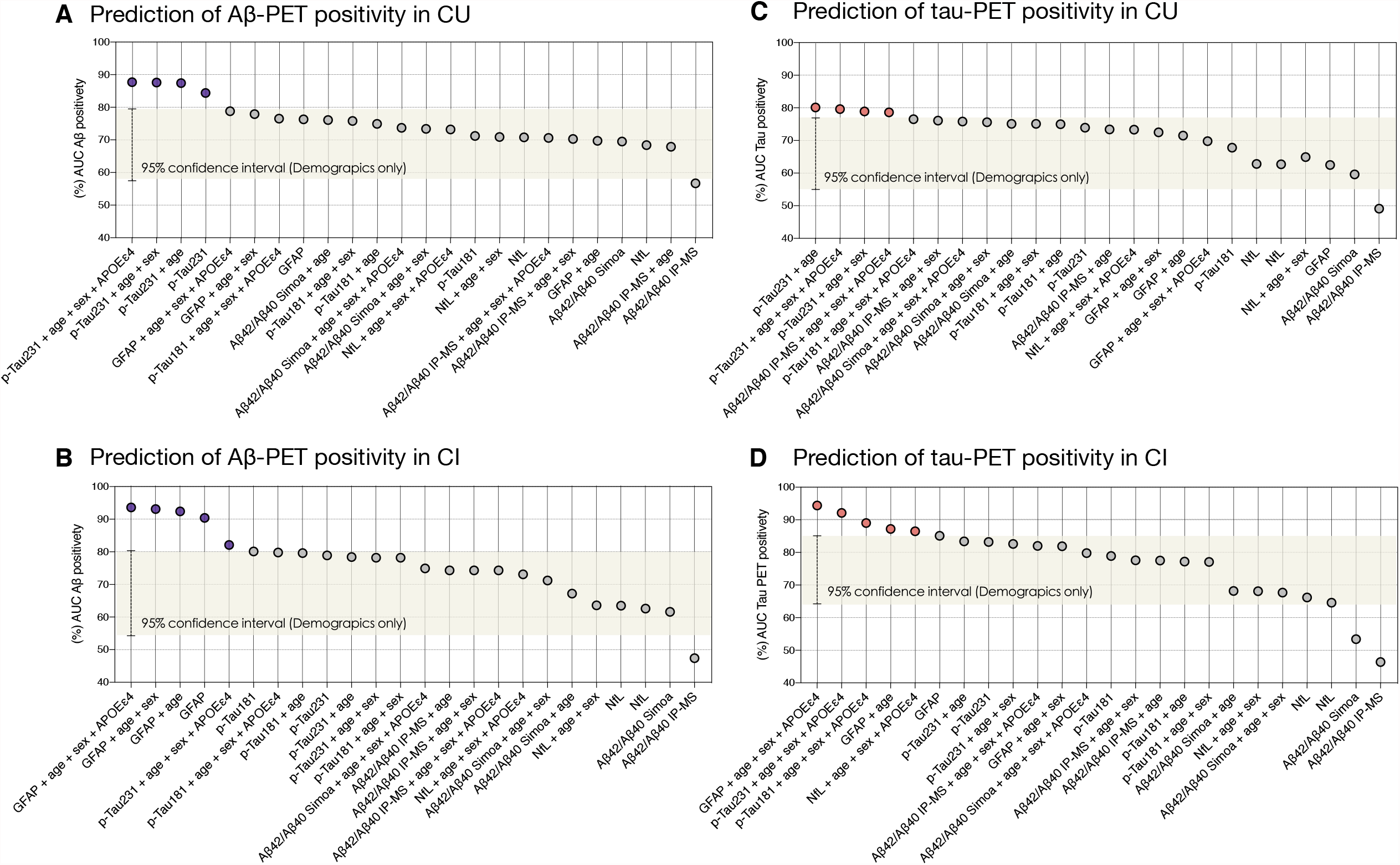
Performance of plasma markers to identify Al3- and Tau-PET positivity. The plots on the left side show the AUC values for the prediction of Al3-PET positivity using plasma biomarkers alone and adjusted for different demographics variable combinations in (A) CU and (B) Cl individuals. The plots on the right side show the AUC values for predicting Tau-PET positivity of plasma biomarkers alone and adjusted for different demographics variable combinations in (C) CU and (D) Cl individuals. The area in the rectangle represents 95% confidence interval of the model using all the demographic variables to predict PET positivity (age, sex, and APOE e4). CU = Cognitively unimpaired, Cl = Cognitively impaired.

### 3.8 Prediction of tau-PET positivity with plasma biomarker concentrations

In the CU group, p-tau231 (alone or accounting for demographic characteristics) and Aβ42/40 (IP-MS) accounting for demographics presented a higher performance in predicting tau-PET positivity than demographics alone (P < 0.05; Figure 4C) to detect tau-PET positivity. In contrast, Aβ42/40 (Simoa), p-tau181, and NfL (alone or accounting for demographic characteristics) did not show a better a better predictive performance than demographics alone (95% confidence interval range: 0.579-0.761). DeLong test comparisons showed that the model including p-tau231 and demographics (AUC = 79.6%) was a significantly better predictor of tau-PET positivity than a model including with Aβ42/40 (measured by Simoa and IP-MS), GFAP, and NfL alone (Table 2) or accounting for demographics (Supplemental Table 3). In the CI group, the models including GFAP, p-tau231, and p-tau181 (alone or accounting for demographic characteristics), NfL accounting for demographics showed a higher performance than demographics alone (P < 0.0001; Figure 4D) to predict tau-PET positivity. In contrast, Aβ42/40 (Simoa and IP-MS) and NfL (alone or accounting for one or two demographics characteristics) did not show a better predictive performance than demographics alone (95% confidence interval range: 0.615-0.867). DeLong test comparisons showed that the model including GFAP plus demographics (AUC = 94.4%) was significantly better in predicting tau-PET positivity than the performance of models including Aβ42/40 (Simoa and IP-MS), p-tau231, p-tau181, and NfL alone (Table 2) or with demographics (Supplemental Table 3).

## 4. Discussion

In this study, we compared plasma Aβ42/40 (Simoa/IP-MS), p-tau, GFAP, and NfL biomarkers in the same individuals to identify Aβ- and tau-PET abnormality in the aging and AD dementia. We demonstrated that plasma p-tau, GFAP, and NfL significantly correlated with each other, while plasma Aβ42/40 (Simoa/IP-MS) did not correlate with the other plasma markers. Our main findings were that plasma p-tau231 showed the best discriminative accuracy to both Aβ- and tau-PET positivity in CU, whereas plasma GFAP had the best discriminative accuracy to both Aβ- and tau-PET positivity in CI. In addition, we also demonstrated that including demographics (age, sex, and APOE ε4) increases the discriminative performance of the plasma markers.

We observed that plasma Aβ42/40 assays measured by both Simoa, and IP-MS did not correlate with other plasma markers and showed only a moderate ability to identify Aβ- and tau-PET positivity in CU and CI individuals. Specifically, we demonstrated that the addition of Aβ42/40 (Simoa and IP-MS) did not increase the predictive performance of demographics (age, sex, and APOE ε4) to identify Aβ-or tau-PET positivity. These results add to the conflicting literature in which some studies suggest that plasma Aβ42 or Aβ42/40 ratio strongly correlated with brain Aβ (AUC = 88 to 97%) [21, 25], while others suggest only a moderate association between plasma and PET assessments of Aβ [43, 44]. Moreover, previous research revealed that IP-MS performed better than Simoa in terms of Aβ estimates [23], whereas we found similar discriminative accuracy using both techniques. Several biological and analytical factors may contribute to the divergence of our results. For example, biological factors such as peripheral Aβ expression accounts to >50% of the global plasma Aβ [45] and the modest fold-change between CU and CI in plasma (10–20%) compared to CSF Aβ (40– 60%) may lead to the high susceptibility of plasma Aβ measures to small variabilities in pre-analytical approaches and cohorts characteristics [20, 21, 44]. In addition, differences in the analytical performance of distinct techniques used to measure plasma Aβ can also play a role in the difference in performance found across studies [23]. We have shown lack of robust correlation between plasma Aβ values and the other AD-related plasma markers. It is important to emphasize that although previous studies have shown a higher performance of plasma Aβ to detect brain Aβ than the presented here, to the best of our knowledge, no previous studies have shown a robust correlation between plasma Aβ and other plasma markers such as p-tau and GFAP. Altogether, these findings suggest that more studies are needed to elucidate the robustness of blood based Aβ assays to identify AD pathophysiology.

We show that plasma GFAP, compared to Aβ, p-tau, and NfL, better discriminates both Aβ- and tau-PET positivity in CI individuals. These results align with recent findings showing that GFAP levels progressively increase with AD progression [10, 16, 17]. Furthermore, the significant positive association between GFAP and Aβ-PET in CU individuals found in our study corroborated recent findings showing that GFAP levels are increased in individuals at risk of AD dementia [10, 46]. Interestingly, in CI group the GFAP levels were also highly associated with tau-PET positivity, independent of Aβ, in our study. Specifically, GFAP outperformed the p-tau markers to predict tau-PET positivity in the CI group. Because it has already been demonstrated that the association of GFAP with tau pathology is mediated by Aβ pathology [10], we speculate that the high performance of GFAP to predict tau-PET positivity is related to the fact that ∼70% of the tau-positive individuals are also Aβ positive. However, we cannot exclude the possibility that GFAP’s performance to identify tau-PET positivity is due to its increase in response to tau tangles deposition [47]. In addition, our findings and results from recent studies showing that plasma GFAP is closely linked to AD pathophysiology [10, 17, 46] suggest that this biomarker has the potential to be used for the categorization of individuals concerning their underlying pathological profiles in biomarker classification methods such as the AT(N) scheme [37, 48].

The study’s main strength is the use of a well-characterized single cohort of individuals that underwent state-of-art harmonized biomarker acquisitions and quantifications However, our findings should be viewed considering some limitations. The cohort used in this study is composed of individuals motivated to participate in a dementia study, potentially being a source of self-selection bias. Moreover, this study includes mostly White individuals, limiting the generalizability of our results to Black and Latinx populations. Previous studies suggest a negative association between Aβ42/40 values and Aβ-PET uptake [49]. Here, we found a positive association between Aβ42/40 IP-MS values and Aβ-PET in CU individuals. Future studies are being planned to elucidate the underpinnings of this unexpected positive association. Although previous studies suggest that plasma p-tau217 assays show better performance to detect AD dementia than p-tau181 and measurements of the plasma p-tau217 epitope were not available in our population. However, we have recently shown that plasma p-tau231 outperforms CSF p-tau217 to identify early changes in Aβ-PET [50].

## 5. Conclusion

To conclude, our results support plasma p-tau231 as an early marker of AD pathophysiology in asymptomatic individuals and GFAP as a robust marker of both Aβ and tau positivity in symptomatic individuals. These results highlight that the performance of plasma biomarkers to detect brain AD pathophysiology is disease stage specific.

## Data Availability

All data produced in the present study are available upon reasonable request to the authors.

## Reference

[1] Jack CR, Jr., Knopman DS, Jagust WJ, Petersen RC, Weiner MW, Aisen PS, et al. Tracking pathophysiological processes in Alzheimer’s disease: an updated hypothetical model of dynamic biomarkers. Lancet Neurol. 2013;12:207–16.

[2] Jack CR, Jr., Bennett DA, Blennow K, Carrillo MC, Dunn B, Haeberlein SB, et al. NIA-AA Research Framework: Toward a biological definition of Alzheimer’s disease. Alzheimers Dement. 2018;14:535–62.

[3] Morris JC, Schindler SE, McCue LM, Moulder KL, Benzinger TLS, Cruchaga C, et al. Assessment of Racial Disparities in Biomarkers for Alzheimer Disease. JAMA Neurol. 2019;76:264–73.

[4] Molinuevo JL, Ayton S, Batrla R, Bednar MM, Bittner T, Cummings J, et al. Current state of Alzheimer’s fluid biomarkers. Acta Neuropathol. 2018;136:821–53.

[5] Klunk WE, Engler H, Nordberg A, Wang Y, Blomqvist G, Holt DP, et al. Imaging brain amyloid in Alzheimer’s disease with Pittsburgh Compound-B. Ann Neurol. 2004;55:306–19.

[6] Villemagne VL, Fodero-Tavoletti MT, Masters CL, Rowe CC. Tau imaging: early progress and future directions. Lancet Neurol. 2015;14:114–24.

[7] Villemagne VL, Doré V, Burnham SC, Masters CL, Rowe CC. Imaging tau and amyloid-β proteinopathies in Alzheimer disease and other conditions. Nat Rev Neurol. 2018;14:225–36.

[8] Karikari TK, Pascoal TA, Ashton NJ, Janelidze S, Benedet AL, Rodriguez JL, et al. Blood phosphorylated tau 181 as a biomarker for Alzheimer’s disease: a diagnostic performance and prediction modelling study using data from four prospective cohorts. Lancet Neurol. 2020;19:422–33.

[9] Karikari TK, Benedet AL, Ashton NJ, Lantero Rodriguez J, Snellman A, Suárez-Calvet M, et al. Diagnostic performance and prediction of clinical progression of plasma phospho-tau181 in the Alzheimer’s Disease Neuroimaging Initiative. Mol Psychiatry. 2021;26:429–42.

[10] Benedet AL, Milà-Alomà M, Vrillon A, Ashton NJ, Pascoal TA, Lussier F, et al. Differences Between Plasma and Cerebrospinal Fluid Glial Fibrillary Acidic Protein Levels Across the Alzheimer Disease Continuum. JAMA Neurology. 2021.

[11] Palmqvist S, Janelidze S, Quiroz YT, Zetterberg H, Lopera F, Stomrud E, et al. Discriminative Accuracy of Plasma Phospho-tau217 for Alzheimer Disease vs Other Neurodegenerative Disorders. Jama. 2020;324:772–81.

[12] Mattsson-Carlgren N, Janelidze S, Palmqvist S, Cullen N, Svenningsson AL, Strandberg O, et al. Longitudinal plasma p-tau217 is increased in early stages of Alzheimer’s disease. Brain. 2020;143:3234–41.

[13] Ashton NJ, Pascoal TA, Karikari TK, Benedet AL, Lantero-Rodriguez J, Brinkmalm G, et al. Plasma p-tau231: a new biomarker for incipient Alzheimer’s disease pathology. Acta Neuropathol. 2021;141:709–24.

[14] Simpson JE, Ince PG, Lace G, Forster G, Shaw PJ, Matthews F, et al. Astrocyte phenotype in relation to Alzheimer-type pathology in the ageing brain. Neurobiol Aging. 2010;31:578–90.

[15] Oeckl P, Halbgebauer S, Anderl-Straub S, Steinacker P, Huss AM, Neugebauer H, et al. Glial Fibrillary Acidic Protein in Serum is Increased in Alzheimer’s Disease and Correlates with Cognitive Impairment. J Alzheimers Dis. 2019;67:481–8.

[16] Pereira JB, Janelidze S, Smith R, Mattsson-Carlgren N, Palmqvist S, Teunissen CE, et al. Plasma GFAP is an early marker of amyloid-β but not tau pathology in Alzheimer’s disease. Brain. 2021.

[17] Cicognola C, Janelidze S, Hertze J, Zetterberg H, Blennow K, Mattsson-Carlgren N, et al. Plasma glial fibrillary acidic protein detects Alzheimer pathology and predicts future conversion to Alzheimer dementia in patients with mild cognitive impairment. Alzheimer’s Research & Therapy. 2021;13:68.

[18] Mattsson N, Andreasson U, Zetterberg H, Blennow K. Association of Plasma Neurofilament Light With Neurodegeneration in Patients With Alzheimer Disease. JAMA Neurol. 2017;74:557–66.

[19] Ashton NJ, Janelidze S, Al Khleifat A, Leuzy A, van der Ende EL, Karikari TK, et al. A multicentre validation study of the diagnostic value of plasma neurofilament light. Nature Communications. 2021;12:3400.

[20] Schindler SE, Bollinger JG, Ovod V, Mawuenyega KG, Li Y, Gordon BA, et al. High-precision plasma β-amyloid 42/40 predicts current and future brain amyloidosis. Neurology. 2019;93:e1647–e59.

[21] Nakamura A, Kaneko N, Villemagne VL, Kato T, Doecke J, Doré V, et al. High performance plasma amyloid-β biomarkers for Alzheimer’s disease. Nature. 2018;554:249–54.

[22] De Meyer S, Schaeverbeke JM, Verberk IMW, Gille B, De Schaepdryver M, Luckett ES, et al. Comparison of ELISA- and SIMOA-based quantification of plasma Aβ ratios for early detection of cerebral amyloidosis. Alzheimers Res Ther. 2020;12:162.

[23] Janelidze S, Teunissen CE, Zetterberg H, Allué JA, Sarasa L, Eichenlaub U, et al. Head-to-Head Comparison of 8 Plasma Amyloid-β 42/40 Assays in Alzheimer Disease. JAMA Neurol. 2021.

[24] Pannee J, Shaw LM, Korecka M, Waligorska T, Teunissen CE, Stoops E, et al. The global Alzheimer’s Association round robin study on plasma amyloid β methods. Alzheimers Dement (Amst). 2021;13:e12242.

[25] Ovod V, Ramsey KN, Mawuenyega KG, Bollinger JG, Hicks T, Schneider T, et al. Amyloid β concentrations and stable isotope labeling kinetics of human plasma specific to central nervous system amyloidosis. Alzheimer’s & dementia : the journal of the Alzheimer’s Association. 2017;13:841–9.

[26] Thijssen EH, Verberk IMW, Vanbrabant J, Koelewijn A, Heijst H, Scheltens P, et al. Highly specific and ultrasensitive plasma test detects Abeta(1-42) and Abeta(1-40) in Alzheimer’s disease. Sci Rep. 2021;11:9736.

[27] Sato C, Barthélemy NR, Mawuenyega KG, Patterson BW, Gordon BA, Jockel-Balsarotti J, et al. Tau Kinetics in Neurons and the Human Central Nervous System. Neuron. 2018;97:1284-98.e7.

[28] Hampel H, O’Bryant SE, Molinuevo JL, Zetterberg H, Masters CL, Lista S, et al. Blood-based biomarkers for Alzheimer disease: mapping the road to the clinic. Nat Rev Neurol. 2018;14:639–52.

[29] Mattsson N, Cullen NC, Andreasson U, Zetterberg H, Blennow K. Association Between Longitudinal Plasma Neurofilament Light and Neurodegeneration in Patients With Alzheimer Disease. JAMA Neurol. 2019;76:791–9.

[30] Hansson O, Janelidze S, Hall S, Magdalinou N, Lees AJ, Andreasson U, et al. Blood-based NfL: A biomarker for differential diagnosis of parkinsonian disorder. Neurology. 2017;88:930–7.

[31] Bridel C, van Wieringen WN, Zetterberg H, Tijms BM, Teunissen CE, Alvarez-Cermeño JC, et al. Diagnostic Value of Cerebrospinal Fluid Neurofilament Light Protein in Neurology: A Systematic Review and Meta-analysis. JAMA Neurol. 2019;76:1035–48.

[32] Chatterjee P, Pedrini S, Stoops E, Goozee K, Villemagne VL, Asih PR, et al. Plasma glial fibrillary acidic protein is elevated in cognitively normal older adults at risk of Alzheimer’s disease. Translational Psychiatry. 2021;11:27.

[33] Medeiros R, LaFerla FM. Astrocytes: conductors of the Alzheimer disease neuroinflammatory symphony. Exp Neurol. 2013;239:133–8.

[34] Therriault J, Benedet AL, Pascoal TA, Savard M, Ashton NJ, Chamoun M, et al. Determining Amyloid-β Positivity Using (18)F-AZD4694 PET Imaging. J Nucl Med. 2021;62:247–52.

[35] McKhann GM, Knopman DS, Chertkow H, Hyman BT, Jack CR, Jr., Kawas CH, et al. The diagnosis of dementia due to Alzheimer’s disease: recommendations from the National Institute on Aging-Alzheimer’s Association workgroups on diagnostic guidelines for Alzheimer’s disease. Alzheimers Dement. 2011;7:263–9.

[36] Pascoal TA, Therriault J, Benedet AL, Savard M, Lussier FZ, Chamoun M, et al. 18F-MK-6240 PET for early and late detection of neurofibrillary tangles. Brain. 2020;143:2818–30.

[37] Jack CR, Jr., Wiste HJ, Weigand SD, Therneau TM, Lowe VJ, Knopman DS, et al. Defining imaging biomarker cut points for brain aging and Alzheimer’s disease. Alzheimers Dement. 2017;13:205–16.

[38] Gisslén M, Price RW, Andreasson U, Norgren N, Nilsson S, Hagberg L, et al. Plasma Concentration of the Neurofilament Light Protein (NFL) is a Biomarker of CNS Injury in HIV Infection: A Cross-Sectional Study. EBioMedicine. 2015;3:135–40.

[39] Mielke MM, Hagen CE, Xu J, Chai X, Vemuri P, Lowe VJ, et al. Plasma phospho-tau181 increases with Alzheimer’s disease clinical severity and is associated with tau- and amyloid-positron emission tomography. Alzheimers Dement. 2018;14:989–97.

[40] Keshavan A, Pannee J, Karikari TK, Rodriguez JL, Ashton NJ, Nicholas JM, et al. Population-based blood screening for preclinical Alzheimer’s disease in a British birth cohort at age 70. Brain. 2021;144:434–49.

[41] Mathotaarachchi S, Wang S, Shin M, Pascoal TA, Benedet AL, Kang MS, et al. VoxelStats: A MATLAB Package for Multi-Modal Voxel-Wise Brain Image Analysis. Front Neuroinform. 2016;10:20.

[42] Larsen WA, McCleary SJ. The Use of Partial Residual Plots in Regression Analysis. Technometrics. 1972;14:781–90.

[43] Verberk IMW, Slot RE, Verfaillie SCJ, Heijst H, Prins ND, van Berckel BNM, et al. Plasma Amyloid as Prescreener for the Earliest Alzheimer Pathological Changes. Ann Neurol. 2018;84:648–58.

[44] Janelidze S, Stomrud E, Palmqvist S, Zetterberg H, van Westen D, Jeromin A, et al. Plasma β-amyloid in Alzheimer’s disease and vascular disease. Sci Rep. 2016;6:26801.

[45] Citron M, Vigo-Pelfrey C, Teplow DB, Miller C, Schenk D, Johnston J, et al. Excessive production of amyloid beta-protein by peripheral cells of symptomatic and presymptomatic patients carrying the Swedish familial Alzheimer disease mutation. Proc Natl Acad Sci U S A. 1994;91:11993–7.

[46] Chatterjee P, Pedrini S, Stoops E, Goozee K, Villemagne VL, Asih PR, et al. Plasma glial fibrillary acidic protein is elevated in cognitively normal older adults at risk of Alzheimer’s disease. Transl Psychiatry. 2021;11:27.

[47] Garwood CJ, Ratcliffe LE, Simpson JE, Heath PR, Ince PG, Wharton SB. Review: Astrocytes in Alzheimer’s disease and other age-associated dementias: a supporting player with a central role. Neuropathol Appl Neurobiol. 2017;43:281–98.

[48] Hampel H, Cummings J, Blennow K, Gao P, Jack CR, Vergallo A. Developing the ATX(N) classification for use across the Alzheimer disease continuum. Nature Reviews Neurology. 2021;17:580–9.

[49] Risacher SL, Fandos N, Romero J, Sherriff I, Pesini P, Saykin AJ, et al. Plasma amyloid beta levels are associated with cerebral amyloid and tau deposition. Alzheimers Dement (Amst). 2019;11:510–9.

[50] Karikari TK, Emeršič A, Vrillon A, Lantero-Rodriguez J, Ashton NJ, Kramberger MG, et al. Head-to-head comparison of clinical performance of CSF phospho-tau T181 and T217 biomarkers for Alzheimer’s disease diagnosis. Alzheimers Dement. 2021;17:755–67.

