## Supplemental Figure 1 for "Performance of plasma amyloid, tau, and astrocyte biomarkers to identify cerebral AD pathophysiology"

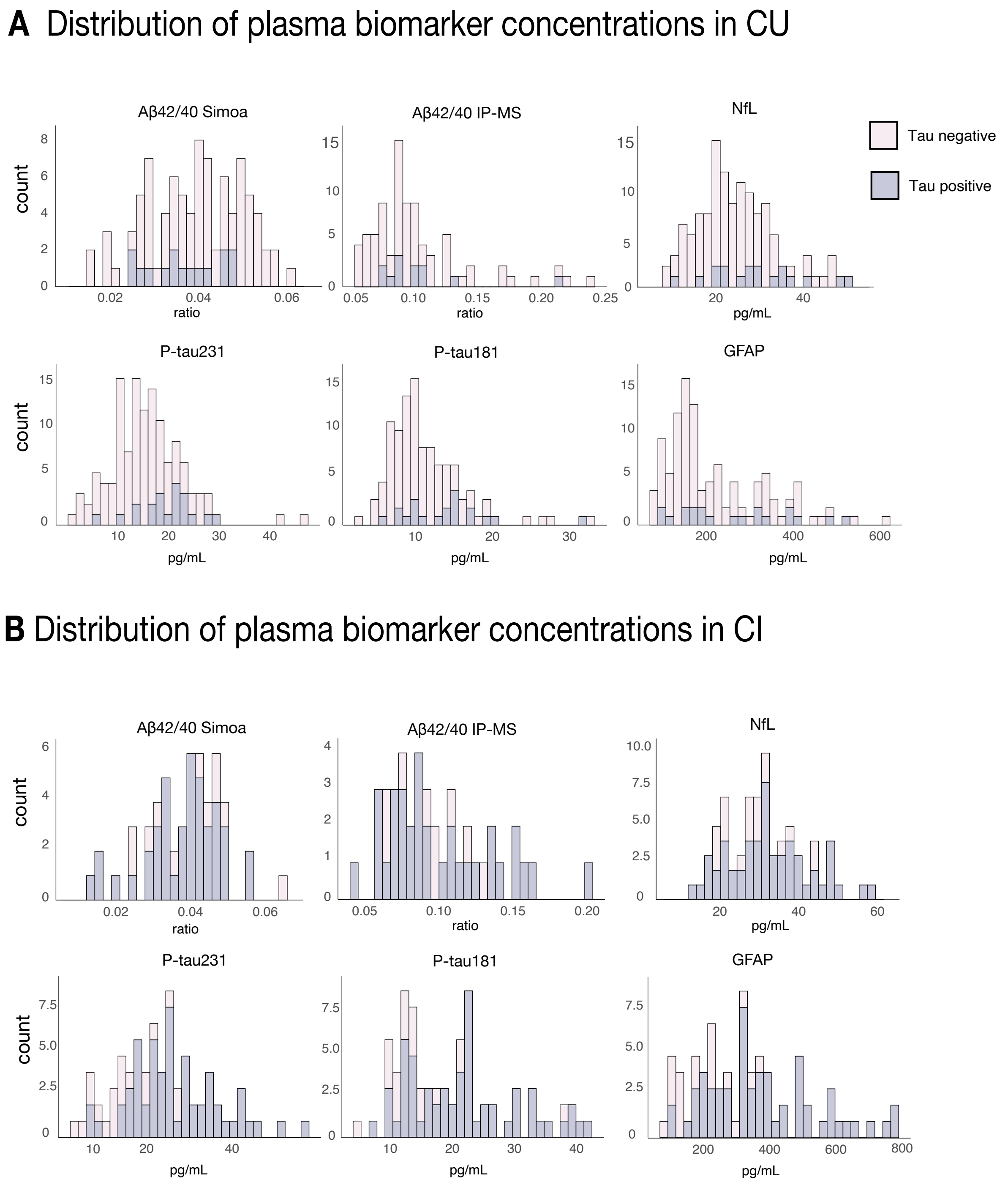


**Supplemental Figure 1.** Distribution of plasma biomarkers in CU and CI individuals. The figure shows the histograms with the distribution of each biomarker regarding their Tau status in CU (A) and CI (B) groups.
