## Supplemental Figure 2 for "Performance of plasma amyloid, tau, and astrocyte biomarkers to identify cerebral AD pathophysiology"

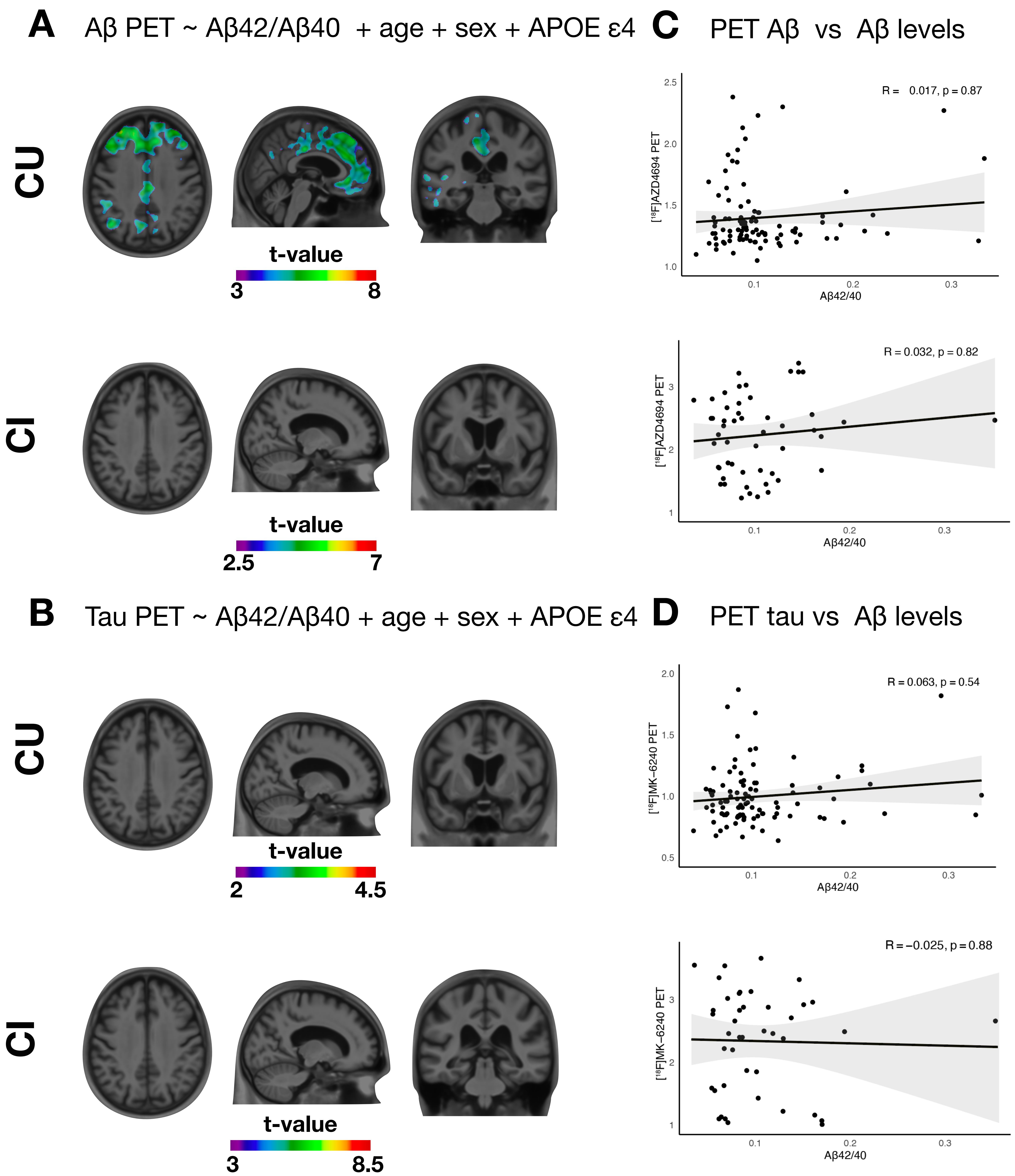


**Supplemental Figure 2. Voxel-wise associations of [18F]AZD4694 SUVR and [18F]MK-6240 SUVR with plasma Aβ42/40 IP-MS.** The figure shows voxel-wise t-statistical maps of linear regressions between PET SUVR and plasma Aβ42/40 IP-MS FDR-corrected for multiple comparisons at $P < 0.05$. Panel **A** shows the regions with a significant positive association between Aβ [18F]AZD4694 and plasma Aβ42/40 IP-MS CU individuals. No negative associations were found after correction for multiple comparisons. The panel **B** shows no significant association between tau [18F]MK-6240 SUVR and plasma Aβ42/40 IP-MS in CU and CI individuals. No negative association was found between biomarkers after multiple comparison correction. In Panel **C,** the scatterplots and Spearman correlations between the Aβ42/40 IP-MS measures and Aβ- PET SUVR showed no significant association between the markers. In Panel **D,** the scatterplots and Spearman correlations between Aβ42/40 IP-MS measures and Tau- PET SUVR showed no significant association between the markers.
