## Supplemental Figure 3 for "Performance of plasma amyloid, tau, and astrocyte biomarkers to identify cerebral AD pathophysiology"

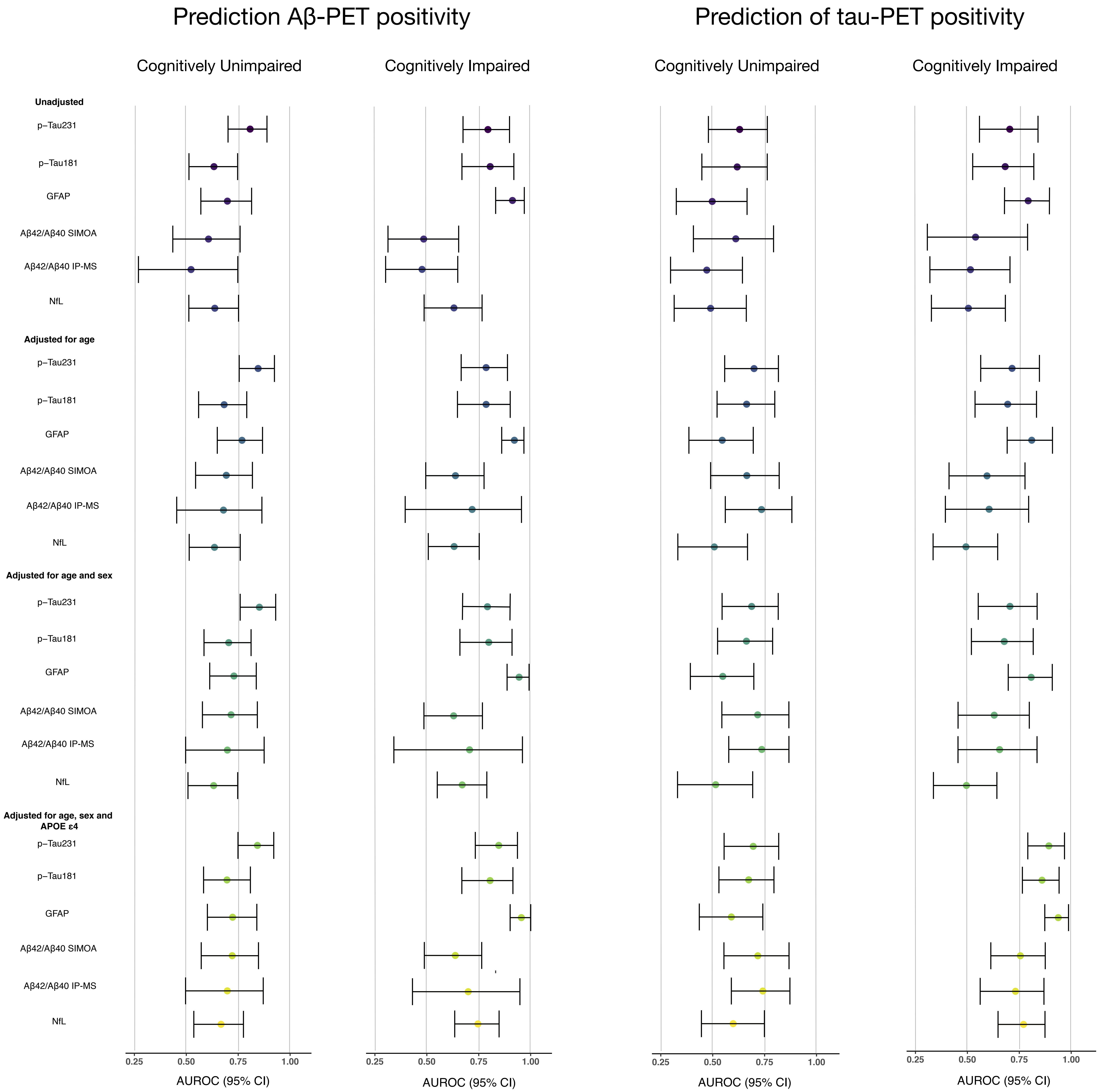


**Supplemental Figure 3.** Association of Aβ- and Tau-PET positivity with plasma biomarker concentrations. The area under the curve with their respective 95% confidence interval for the prediction of Aβ- and Tau-PET positivity for each plasma biomarkers alone or in combination with sec, age, and APOE ε4.
