## Supplemental Table 1 for "Performance of plasma amyloid, tau, and astrocyte biomarkers to identify cerebral AD pathophysiology"

**Supplementary Table 1.** **Associations of plasma biomarkers with Aβ-PET and tau-PET.**

| | CU | | | | CI | | | |
|---|---|---|---|---|---|---|---|---|
| | **β (95% Confidence interval)** | ***T*-value** | ***P*-value** | **R-squared[a]** | **β (95% Confidence interval)** | ***T*-value** | ***P*-value** | **R-squared[a]** |
| **Aβ PET SUVR ~ Plasma biomarker concentration + age + sex + APOE ε4 status** | | | | | | | | |
| Aβ42/40 Simoa | -5.67 (-0.33 to 0.008) | -1.4 | 0.053 | 0.18 | -0.16 (-0.76 to 0.86) | -1.7 | 0.177 | 0.16 |
| Aβ42/40 IP-MS | 0.11 (-0.06 to 0.28) | 1.3 | 0.202 | 0.07 | -0.31 (-1.47 to 0.84) | -0.6 | 0.583 | 0.15 |
| p-tau231 | 0.49 (0.02 to 0.03) | 6.8 | < 0.001* | 0.33 | 0.49 (0.32 to 0.68) | 5.4 | < 0.001* | 0.30 |
| p-tau181 | 0.25 (0.08 to 0.41) | 2.9 | 0.003* | 0.16 | 0.33 (0.13 to 0.53) | 3.3 | < 0.001* | 0.16 |
| GFAP | 0.28 (-0.09 to 0.47) | 2.9 | 0.004* | 0.16 | 0.49 (-0.29 to 0.67) | 5.2 | < 0.001* | 0.28 |
| NfL | 0.08 (-0.18 to 0.35) | 0.6 | 0.522 | 0.10 | 0.06 (-0.16 to 0.29) | 0.7 | 0.576 | 0.13 |
| **Tau PET SUVR ~ Plasma biomarker concentration + age + sex + APOE ε4 status** | | | | | | | | |
| Aβ42/40 Simoa | -3.80 (-7.87 to 0.27) | -1.8 | 0.07 | 0.10 | -0.06 (-0.30 to 0.19) | -0.5 | 0.633 | 0.23 |
| Aβ42/40 IP-MS | 0.12 (-0.05 to 0.30) | 1.4 | 0.172 | 0.08 | -0.03 (-0.32 to 0.25) | -0.2 | 0.818 | 0.21 |
| p-tau231 | 0.23 (0.07 to 0.39) | 2.8 | 0.003 | 0.15 | 0.51 (0.34 to 0.676) | 6.1 | < 0.001* | 0.45 |
| p-tau181 | 0.22 (0.06 to 0.39) | 2.7 | 0.007 | 0.15 | 0.42 (0.25 to 0.59) | 4.9 | < 0.001* | 0.37 |
| GFAP | 0.06 (-0.14 to 0.25) | 0.5 | 0.568 | 0.10 | 0.50 (0.33 to 0.67) | 5.9 | < 0.001* | 0.44 |
| NfL | 0.003 (-0.17 to 0.18) | 0.03 | 0.977 | 0.07 | 0.35 (0.13 to 0.56) | 3.3 | 0.001* | 0.36 |

Aβ = amyloid-β; p-tau181 = tau phosphorylated at threonine 181; p-tau231 = tau phosphorylated at threonine 231; GFAP = glial fibrillary acidic protein, NfL = neurofilament light; PET = positron emission tomography; SUVR = standardized uptake value ratio; [a]Adjusted R-squared. * Indicates that correlations the survived to Bonferroni correction for multiple comparison (12 tests, corrected P value $< 0.0042$).
