## Supplemental Table 2 for "Performance of plasma amyloid, tau, and astrocyte biomarkers to identify cerebral AD pathophysiology"

**Supplementary Table 2. Associations of plasma biomarkers with Aβ-PET and tau-PET.**

| | CU | | | CI | | |
|---|---|---|---|---|---|---|
| | **β (95% Confidence interval)** | ***T*-value** | ***P*-value** | **β (95% Confidence interval)** | ***T*-value** | ***P*-value** |
| **Aβ PET SUVR ~ Plasma Aβ42/40 Simoa + Tau PET SUVR + age + sex + APOE ε4 status** | | | | | | |
| Aβ42/40 Simoa | -4.4 (-7.7 to -1.03) | 2.6 | 0.011 | -0.17 (-0.34 to 0.07) | -1.3 | 0.195 |
| Tau PET SUVR | 0.35 (0.18 to 0.51) | 4.2 | < 0.001* | 0.56 (-0.33 to 0.78) | 4.9 | < 0.001* |
| **Aβ PET SUVR ~ Plasma Aβ42/40 IP-MS + Tau PET SUVR + age + sex + APOE ε4 status** | | | | | | |
| Aβ42/40 IP-MS | 0.07 (-0.09 to 0.23) | 0.8 | 0.424 | 0.11 (-0.12 to 0.34) | 0.9 | 0.344 |
| Tau PET SUVR | 0.32 (0.13 to 0.51) | 3.3 | 0.001* | 0.51 (0.26 to 0.75) | 4.1 | < 0.001* |
| **Aβ PET SUVR ~ Plasma p-tau231 concentration + Tau PET SUVR + age + sex + APOE ε4 status** | | | | | | |
| p-tau231 | 0.39 (0.28 to 0.53) | 6.0 | < 0.001* | 0.26 (-0.05 to 0.46) | 2.5 | 0.016 |
| Tau PET SUVR | 0.38 (0.24 to 0.52) | 5.5 | < 0.001* | 0.51 (0.29 to 0.73) | 4.5 | < 0.001* |
| **Aβ PET SUVR ~ Plasma p-tau181 concentration + Tau PET SUVR + age + sex + APOE ε4 status** | | | | | | |
| p-tau181 | 0.14 (-0.01 to 0.29) | 1.8 | 0.07 | 0.08 (-0.11 to 0.27) | 0.83 | 0.411 |
| Tau PET SUVR | 0.44 (0.29 to 0.59) | 5.8 | < 0.001* | 0.62 (0.40 to 0.83) | 5.7 | < 0.001* |
| **Aβ PET SUVR ~ Plasma GFAP concentration + Tau PET SUVR + age + sex + APOE ε4 status** | | | | | | |
| GFAP | 0.26 (0.09 to 0.42) | 3.1 | 0.003* | 0.23 (0.03 to 0.43) | 2.3 | 0.025 |
| Tau PET SUVR | 0.47 (0.32 to 0.68) | 6.5 | < 0.001* | 0.51 (0.30 to 0.73) | 4.7 | < 0.001* |
| **Aβ PET SUVR ~ Plasma NfL concentration + Tau PET SUVR + age + sex + APOE ε4 status** | | | | | | |
| NfL | 0.08 (-0.16 to 0.33) | 0.7 | 0.489 | 0.16 (0.37 to 0.05) | 1.5 | 0.132 |
| Tau PET SUVR | 0.390 (0.22 to 0.56) | 4.6 | < 0.001* | 0.64 (0.40 to 0.89) | 5.2 | < 0.001* |

| | | | | | | |
|---|---|---|---|---|---|---|
| **Tau PET SUVR ~ Plasma Aβ42/40 Simoa + Aβ PET SUVR + age + sex + APOE ε4 status** | | | | | | |
| Aβ42/40 Simoa | -1.19 (-5.08 to 2.7) | -0.6 | 0.546 | -0.13 (-0.33 to 0.07) | -1.3 | 0.195 |
| Aβ PET SUVR | - 0.45 (0.24 to 0.66) | 4.2 | < 0.001* | 0.55 (0.32 to 0.77) | 4.9 | < 0.001* |
| **Tau PET SUVR ~ Plasma Aβ42/40 IP-MS + Aβ PET SUVR + age + sex + APOE ε4 status** | | | | | | |
| Aβ42/40 IP-MS | 0.08 (-0.09 to 0.25) | 0.9 | 0.329 | 0.13 (-0.09 to 0.35) | 1.2 | 0.265 |
| Aβ PET SUVR | 0.37 (0.15 to 0.58) | 3.4 | < 0.001 | 0.52 (0.28 to 0.76) | 4.4 | < 0.001* |
| **Tau PET SUVR ~ Plasma p-tau231 concentration + Aβ PET SUVR + age + sex + APOE ε4 status** | | | | | | |
| p-tau231 | -0.01 (-0.18 to 0.16) | -0.11 | 0.910 | 0.25 (0.05 to 0.45) | 2.5 | 0.02 |
| Aβ PET SUVR | 0.51 (0.32 to 0.05) | 5.5 | < 0.001* | 0.49 (0.28 to 0.72) | 5.5 | < 0.001* |
| **Tau PET SUVR ~ Plasma p-tau181 concentration + Aβ PET SUVR + age + sex + APOE ε4 status** | | | | | | |
| p-tau181 | 0.12 (-0.03 to 0.27) | 1.5 | 0.127 | 0.08 (-0.11 to 0.27) | 0.8 | 0.411 |
| Aβ PET SUVR | 0.47 (0.32 to 0.62) | 5.9 | < 0.001* | 0.61 (0.39 to 0.82) | 5.7 | < 0.001* |
| **Tau PET SUVR ~ Plasma GFAP concentration + Aβ PET SUVR + age + sex + APOE ε4 status** | | | | | | |
| GFAP | -0.09 (-0.27 to 0.09) | -0.9 | 0.326 | 0.30 (0.13 to 0.47) | 3.5 | < 0.001* |
| Aβ PET SUVR | 0.52 (0.35 to 0.68) | 6.5 | < 0.001* | 0.41 (0.24 to 0.59) | 5.5 | < 0.001* |
| **Tau PET SUVR ~ Plasma NfL concentration + Aβ PET SUVR + age + sex + APOE ε4 status** | | | | | | |
| NfL | -0.03 (-0.31 to 0.23) | -0.3 | 0.787 | -0.16 (-0.36 to 0.05) | -1.5 | 0.132 |
| Aβ PET SUVR | 0.47 (0.27 to 0.67) | 4.6 | < 0.001* | 0.63 (0.39 to 0.88) | 5.2 | < 0.001* |

Aβ = amyloid-β; p-tau181 = tau phosphorylated at threonine 181; p-tau231 = tau phosphorylated at threonine 231; GFAP = glial fibrillary acidic protein, NfL = neurofilament light; PET = positron emission tomography; SUVR = standardized uptake value ratio; [a]Adjusted R-squared. * Indicates that correlations the survived to Bonferroni correction for multiple comparison (12 tests, corrected P value < 0.0042).
