## Supplemental Table 3 for "Performance of plasma amyloid, tau, and astrocyte biomarkers to identify cerebral AD pathophysiology"

**Supplementary Table 3**. Performance of plasma biomarker to predict Aβ-PET and tau-PET positivity.

| | **Predict Aβ-PET positivity** | | **Predict Tau-PET positivity** | |
|---|---|---|---|---|
| | **CU** | **CI** | **CU** | **CI** |
| ***Biomarker*** | ***AUC*** | **AUC** | **AUC** | **AUC** |
| **Unadjusted** | | | | |
| Aβ42/Aβ40 Simoa | 0.695[f,g,h] | 0.616[i,m,n] | 0.596[g,i,j,k] | 0.534[c,f,g,h,i,m,n,o,q,r] |
| Aβ42/Aβ40 IP-MS | 0.567[f,g,h] | 0.474[a,b,c,d,e,f,g,h,i,j,k,lm,n,p,r] | 0.491[a,b,cd,eg,h,i,j,k,l,m,n,j,k,l,m,n,o,p,q,r] | 0.464[c,e,f,g,h,i,j,k,l,m,n,o,q,r] |
| p-Tau231 | 0.844[a,d,o,p,q] | 0.789[i,m,n] | 0.739 | 0.832[o,p,q,r] |
| p-Tau181 | 0.712[f,g,h] | 0.801[i,m,n] | 0.678 | 0.789[i,l,o] |
| GFAP | 0.763[f,g,h] | 0.904[a,b,c,g,h,p,q,r] | 0.625[l,g,h,i,j,k] | 0.851 |
| NfL | 0.684[f,g,h] | 0.626[i,m,n,o,] | 0.627 | 0.646[g,h,i,l,m,n,o] |
| **Adjusted for age** | | | | |
| Aβ42/Aβ40 Simoa | 0.761[l,m,n] | 0.672[i,m,n] | 0.751 | 0.682[i,l,m,q,r] |
| Aβ42/Aβ40 IP-MS | 0.679[l,m,n] | 0.743[i,m,n] | 0.734 | 0.775[m,n] |
| p-Tau231 | 0.874 [a,b,c,d,e,f,i,j,k,l,m,n,o,p,q,r] | 0.784[i,m,n] | 0.801[l,m] | 0.834[o,p,q] |
| p-Tau181 | 0.749[l,m,n] | 0.796[i,m,n] | 0.750 | 0.772[i,l,o] |
| GFAP | 0.697[l,m,n] | 0.924[a,b,c,d,e,f,g,h,j,k,l,p,q,r] | 0.715[g,i] | 0.872[p,q] |
| NfL | 0.708[l,m,n] | 0.635[i,m,n,o,] | 0.628 | 0.662[g,h,i,l,m,n,o,p,r] |
| **Adjusted for age, sex** | | | | |
| Aβ42/Aβ40 Simoa | 0.734[f,g,h] | 0.712[m,n,o,] | 0.756 | 0.677[i,l,m,n,o] |
| Aβ42/Aβ40 IP-MS | 0.703[f,g,h] | 0.743[e,m,n,o,] | 0.761 | 0.776 |
| p-Tau231 | 0.876 [a,b,c,d,e,f,i,j,k,l,m,n,o,p,q,r] | 0.782[m,n,i,] | 0.789[l,m] | 0.826[i,o,p,q] |
| p-Tau181 | 0.758[f,g,h] | 0.782[m,n,i,] | 0.751 | 0.771[i,o] |
| GFAP | 0.779[f,g,h] | 0.931[a,b,c,d,e,f,g,h,j,k,l,p,q,r] | 0.725[g] | 0.819[p,q] |

| | | | | |
|---|---|---|---|---|
| NfL | 0.709[f,g,h] | 0.636[i,m,n,o,] | 0.649 | 0.681[i,l,m,n,o,r] |
| **Adjusted for age, sex, APOE ε4 status** | | | | |
| Aβ42/Aβ40 Simoa | 0.737[f,g,h] | 0.749[m,n,o,] | 0.758 | 0.798[o,p] |
| Aβ42/Aβ40 IP-MS | 0.706[f,g,h] | 0.743[m,n,o,] | 0.786 | 0.82[p] |
| p-Tau231 | 0.877 [a,b,c,d,e,f,i,j,l,k,m,n,o,p,q,r] | 0.821[i,p,q,r] | 0.796[l,m] | 0.921[a,b,g,h,j,k,p,q] |
| p-Tau181 | 0.765[f,g,h] | 0.798 [m,n,i,] | 0.765 | 0.890[a,b,j,k,p] |
| GFAP | 0.788[f,g,h] | 0.936[a,b,c,d,e,f,g,h,k,l,p,q,r] | 0.698[g,i] | 0.944[a,b,g,h,j,k,o,p,q] |
| NfL | 0.732[f,g,h] | 0.731[m,n,o,] | 0.733 | 0.865[p,q] |

DeLong test provided significant differences between group: *a* compared to Aβ42/Aβ40 Simoa + age; *b* compared to Aβ42/Aβ40 Simoa + age + sex; *c* compared to Aβ42/Aβ40 Simoa + age + *APOE ε4*; *d* compared to Aβ42/Aβ40 Simoa + age; *e* compared to Aβ42/Aβ40 Simoa + age + sex; *f* compared to Aβ42/Aβ40 Simoa + age + *APOE ε4*; *g* compared to p-tau231 + age; *h* compared to p-tau231 + age + sex; *i* compared to p-tau231 + age + *APOE ε4* ; *j* compared to p-tau181 + age; *k* compared to p-tau181 + age + sex; *l* compared to p-tau181 + age + *APOE ε4* ; *m* compared to GFAP + age; *n* compared to GFAP + age + sex; *o* compared to GFAP + age + *APOE ε4* ; *p* compared to NfL + age; *q* compared to NfL + age + sex; *r* compared to NfL + age + *APOE ε4* .
